# Effect of Adjunctive Prefronto-Cerebellar Transcranial Direct Current Stimulation(tDCS) on Craving and Relapse Prevention in Patients with Alcohol Dependence Syndrome (ADS): A Randomized Sham Controlled Study Protocol

**DOI:** 10.1101/2022.08.05.22278474

**Authors:** Susmita Sarkar, Lokesh Kumar Singh, Ajay Kumar, Sai Krishna Tikka

**Author notes:** **Corresponding Author:** Dr. Lokesh Kumar Singh, Additional Professor, Department of Psychiatry, All India Institute of Medical Sciences, Raipur. Contact no-0091-9827171148.

## Abstract

**BACKGROUND:** There is growing evidence that tDCS in the Dorsolateral Prefrontal Cortex (DLPFC) technique is effective in controlling craving and relapse in ADS patients. Till now the results are inconclusive. Interestingly, the cerebellum is related to impulsive and compulsive behavior which is supported by both structural and functional neuroimaging findings. There are few clinical trials regarding the effect of cerebellar tDCS in ADS.

**OBJECTIVE:** To assess the effect of Prefronto-cerebellar tDCS in reducing craving and relapses in ADS patients.

**METHODS:** The study population will be randomly assigned to either of two groups i.e. Active and Sham tDCS group. The active group will receive 10 sessions over 5 days; 2mA current for 15 minutes. The site of anodal stimulation is 2cm below the Iz (inion) and cathodal stimulation at F3 (left prefrontal) and in the sham group will receive the same except that intensity of the current is 0.2mA. Patients will be assessed for craving at 3-time points.

**CONCLUSION:** It will be helpful in analyzing the effect of stimulating DLPFC as well as the cerebellum which is a potential novel site in reducing craving and relapse in ADS.

**HIGHLIGHTS:** tDCS in ADS is being tried for the last decade mainly focusing on the frontal area. With the current understanding of the role of the cerebellum in regulating impulsive and compulsive behavior, we intend to add the cerebellum as a potential site for neuromodulation in reducing craving and relapse in ADS patients.

## INTRODUCTION

Alcohol use in dependence pattern is a global problem, affecting individuals in almost every domain with its impact on family, society, and as a whole nation. Worldwide 3 million deaths every year result from harmful use of alcohol, this represents 5.3% of all deaths ^1^. Craving (an urge or conscious desire to take the drug) is one of the core features of dependence (among the six criteria of dependence according to ICD-10)^2^. Craving is an important predictor of drug-seeking behavior so as relapse. Hence the reduction of craving is the real challenge in Alcohol Dependence Syndrome (ADS). Current Food and Drug Administration approved treatment modalities for the management of craving include drugs like Naltrexone, Acamprosate. But success rate with these treatments is not satisfactory (prevention of relapse only 40-50%) ^3^. So novel treatment options are in high demand.

For the past few years, there is growing interest in non-invasive brain stimulation (NIBS) as a novel treatment option for substance-use disorders (SUDs). Transcranial direct current stimulation (tDCS) has been the method of choice in this area because of its low cost, ease of use, safety, and portability in comparison with Transcranial Magnetic Stimulation^4^. Recent momentum stems from an understanding of the link between addiction and the neuroscience behind it. The primary hypothesized mechanism of tDCS is neuromodulation leading to long-term potentiation (LTP)- or long-term depression (LTD)-like changes in the synaptic coupling of neurons which is an important mechanism for addiction^5^. Craving has been associated with brain areas related to reward, motivation, and memory, including the Prefrontal cortical area, the striatum and ventral pallidum, mesolimbic dopamine structures, such as the ventral tegmental area, and reward memory areas like the amygdala or the hippocampus, and interestingly the cerebellum. It is hypothesized that there is a reciprocal connection between the cerebellum & prefrontal cortex and also with the ventral tegmental area^6^. Prefronto-cerebellar pathology is established in mental disorders in which compulsive and impulsive phenotypes are common. It is expected that stimulation of cerebellar activity will improve prefrontal functionality and reduce compulsive and impulsive behavior. There is quite an appreciable number of Randomized Control trials that showed promising results on prefrontal tDCS but interventional studies regarding the effect of the cerebellum in reduction of craving are still lacking. Therefore, we are including the cerebellum as a novel site along with DLPFC for neuromodulation by tDCS to analyze its effect on craving as well as relapse.

## METHODOLOGY

### Study design

It will be a Randomized Sham Controlled trial.

### Study settings

The study began on October 2021 at a Government-funded multi-specialty hospital in Raipur, Chhattisgarh, India after completing trial registration in CTRI. The study is designed to be completed one and a half years from the start of the project.

### Study Participants

Patients diagnosed with alcohol dependence syndrome as per ICD-10 DCR^6^ with Clinical Institute Withdrawal Assessment for Alcohol Scale Revised CIWA-Ar score<20 with age between 18 -50 years irrespective of gender will be recruited. Patients with comorbid dependence syndrome of other substances (excluding nicotine and caffeine);any other comorbid psychiatric disorder; alcohol or other substance-related psychotic disorder; alcohol-related complicated withdrawal (delirium tremens, withdrawal seizure); with metal in cranium, pacemaker, history of significant traumatic brain injury; history of epilepsy and Cerebrovascular Accident (CVA); pregnancy; skin problems, such as dermatitis, psoriasis, or eczema over the scalp or if there was any serious adverse event following transcranial electric or magnetic stimulation in the past will be excluded from the study.

### Sample Size

The sample size calculation was made using G*Power Software. The total sample size estimated was 52 [α=0.05; Power (1-β) = 0.9; Number of groups= 2; Number of measurements= 3, Correlation between dependent measures: 0.5, Effect Size: f=0.208 With an expected dropout rate of 20 % (i.e., approximately11 subjects), we intend to enroll a total of 64 participants (32 in each of the active and sham groups).

### Recruitment of participants

Patients who are admitted with alcohol dependence syndrome in the department of psychiatry will be screened for participation in the study. An information sheet providing the details of the study will be provided to patients. Patients will be free to participate or to withdraw from the study at any point in time without any reason. Written informed consent will be taken and included in the study.

### Randomization details

The participants will be randomized into two groups (active tDCS and Sham tDCS) by block randomization in blocks of 4. A person who is not involved in the study will perform the random number allocation. Allocation concealment will be done using sealed opaque envelopes. The Rater will be blind to allocation. The session is being given by the Technician.Recruitment was started only after approval by Institute Ethics Committee (Registration No: ECR/714/Inst/CT/2015/RR-21) and registration in the Clinical Trial Register India (CTRI) [Registration No: CTRI/2021/09/036989. Reference no: REF/2021/05/043836]

### Intervention Details

The Active tDCS group will receive 10 sessions (2 per day) over 5 days; 2mA current and for 15 minutes in each session that is spaced over at least 4 hours. The site of Anodal stimulation is 2cm below the Iz (inion), identified according to the international 10-20 system and cathodal stimulation at F3 (left prefrontal). The <u>Sham tDCS group</u> all parameters same except that intensity of current being 0.2mA. To date, the use of conventional tDCS protocols in human trials (≤40 min, ≤4 mA, ≤7.2 Coulombs) has not produced any reports of a Serious Adverse Effect or irreversible injury^7^. High Definition tDCS (HD-tDCS) Instrument that we are using is STARSTIM-8 (Neuroelectrics Inc, Spain) with 8 channel tES stimulatorSTARSTIM-8 wearable cap with multiple channels is being used. (Fig-2)

**Figure.1.**
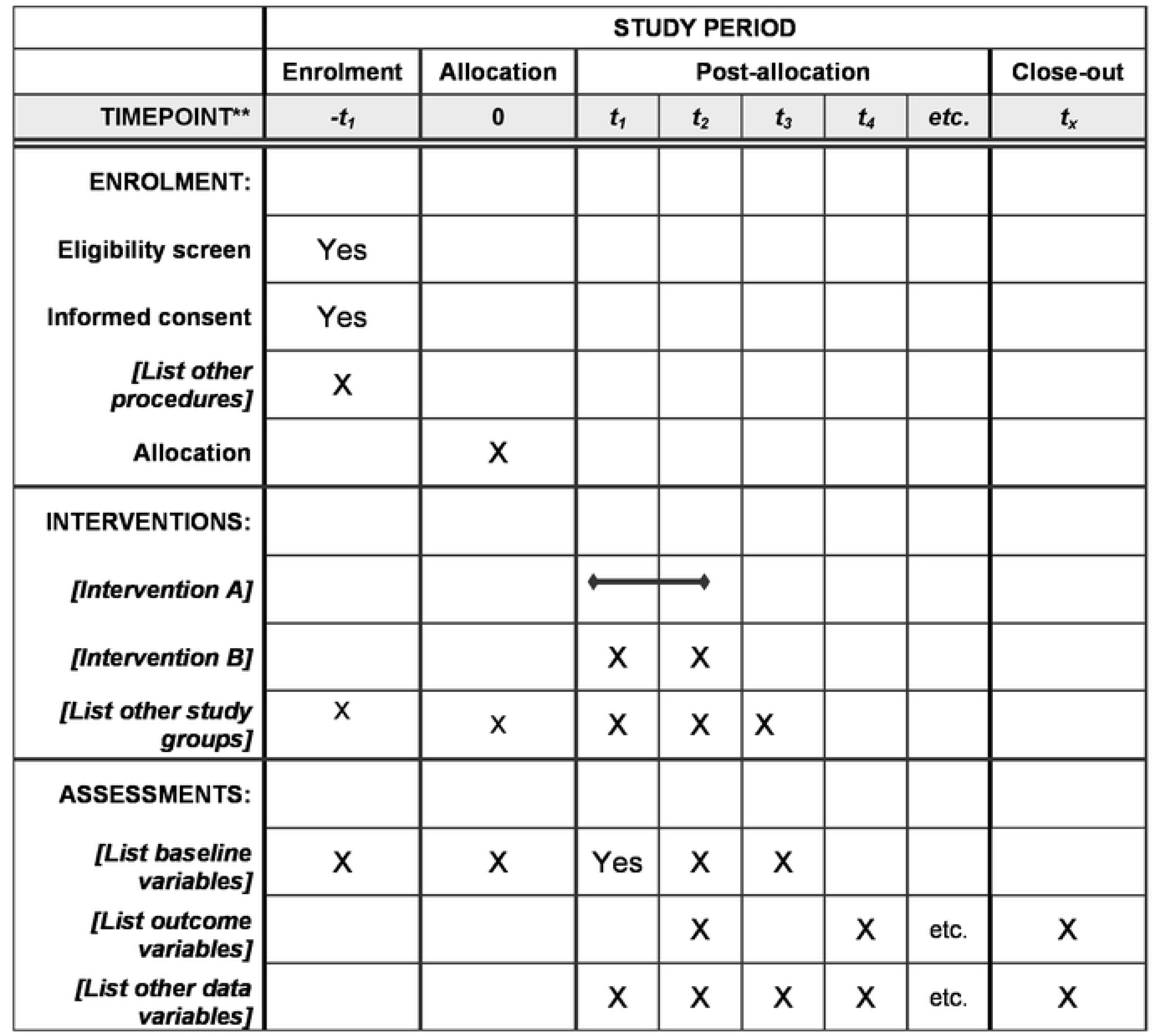
*Abreviations: t1-Before starting tDCS(Day-1) t2-After completion of 10 sessions of tDCS (Day-6) t3-6weeks after the Day of the last tDCS session (Follow up) Intervention A- received Active tDCS Intervention B- received Sham tDCS

**Fig 2.**
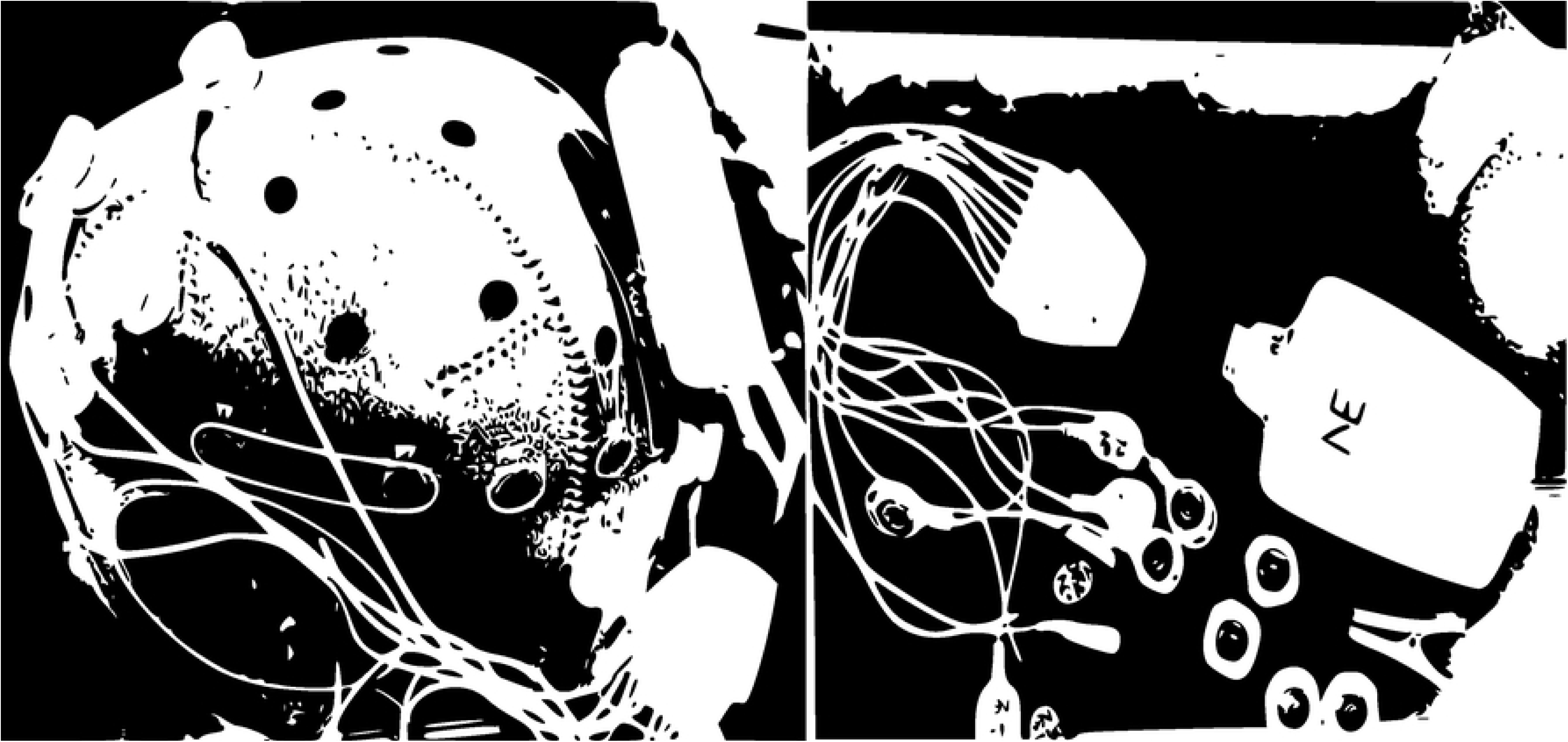
Fig 1 Showing Electrode Placements

### Assessment

The severity of alcohol dependence and nicotine dependence (in a subset of the sample) will be assessed using the Severity of Alcohol Dependence Questionnaire (SADQ) and Fagerstrom Nicotine Dependence Scale (FTND) & Fagerstrom Nicotine Dependence Scale Smokeless Tobacco. The craving will be assessed by ACQ -SF-R (Alcohol Craving Questionaries-Short Form-Revised) Visual Analogue Scale (VAS), Tobacco Craving Questionnaire-Short Form (TCQ-SF) during all 3 time-points. Patients will be assessed for craving at 3-time points; Baseline: Just before the delivery of the first session at around 8 am (Day 1); After completion of 10 sessions of tDCS. On Day 6, 8 am & Follow-up: After 6 weeks from the day of the last session (either face to face or telephonically) for craving thought appropriate scales and relapse.

### Statistical Analysis

The assumption of normality will be verified by normal probability plots and the Shapiro-Wilk test. Group differences for sample characteristics will be examined with an independent t-test and Chi-square test (wherever applicable). An intention-to-treat (ITT) analysis will be conducted to include all the patients recruited in the study, irrespective of whether they completed the study period or not. The missing values will be replaced using the last observation carried forward (LOCF) method. The main analysis will be the interaction effect of treatment overtime (across the three-time points—baseline, completion of the 10th tDCS session, i.e., 6th day and the 30th day) and group (between ‘active’ and sham groups) in the double-blind phase. The overall effect of treatment over time for the two groups will be analyzed using the restricted maximum likelihood (ReML) mixed (growth curve) model analysis with allocation order as ‘subjects’, treatment (active/sham) as the between-subject factor, and time (baseline, completion of 10th tDCS session, follow-up) as the within-subject factor. A supplementary per-protocol (PP)analysis including subjects who have completed the study will also be conducted.

## FLOW CHART OF WORK

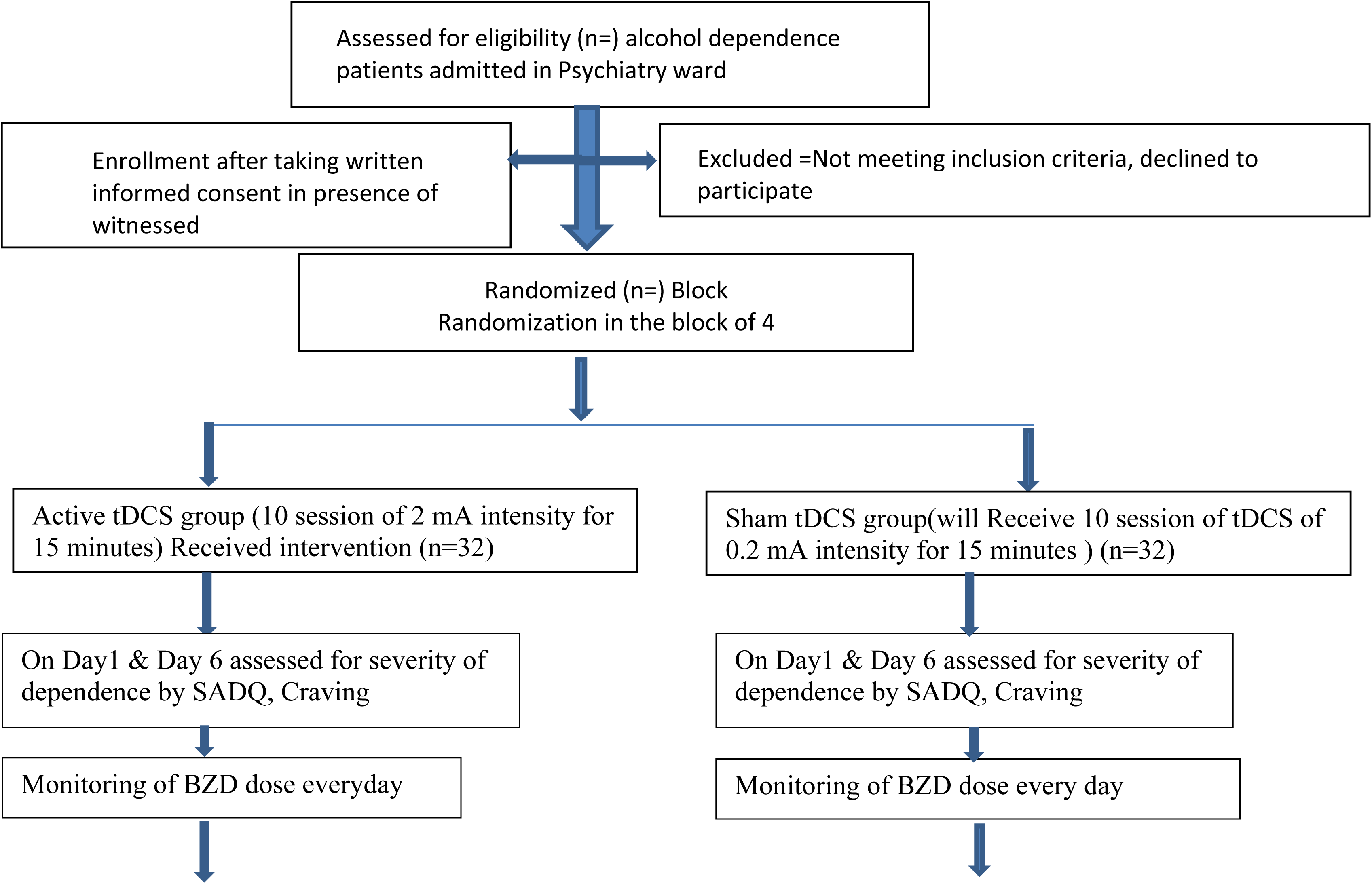

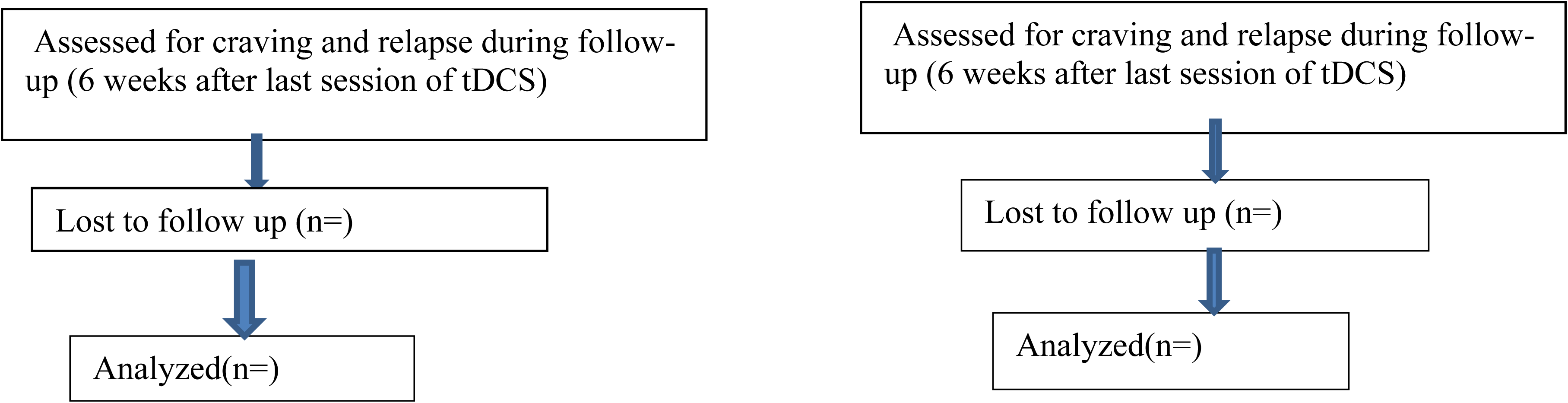

## DISCUSSION

The study of addiction and impulse control disorders has shown that behaviors of seeking and consumption of addictive substances are due to neurobiological alterations which are specifically related to brain networks for reward, stress, and executive control, representing the brain’s adaptation to the continued use of an addictive substance. Dopaminergic circuitry mainly in frontal brain areas is responsible for changes in motivation, attention, habits, and executive control^5^. In this context, tDCS has been proposed as a cheap, safe, and easy way to modulate frontal brain areas and circuitries involved in executive control modulating automatic responses related to impulse control, craving & providing appropriate tools to change brain activity in targeted brain areas and mould neuroplasticity at the network level. The brain areas involved in addiction are the Prefrontal cortex, the striatum and ventral pallidum, mesolimbic dopamine structures, such as the ventral tegmental area, and memory areas like the amygdala or the hippocampus, and interestingly the cerebellum. The cerebellum is related to impulsive and compulsive behavior which is supported by both structural and functional neuroimaging findings (decreased GM volume in several regions of the cerebellum and increased basal ganglia-cerebellar connectivity in patients with the compulsive disorder). Importantly, there are cerebellar activations when drug-related cues are presented which is supported by neuroimaging studies of cue reactivity in drug addicts. It is hypothesized that the cerebellum, by activating drug-cue representations during abstinence, can contribute to compulsive drug-seeking driven by both negative and positive reinforcement^6^. Response inhibition (an executive function of the cerebellum, refers to the ability to suppress inappropriate but prepotent responses) can be improved by stimulating the crebellum^8^. Kim, et.al 2020 in a systemic review and meta-analysis study analyzed a total of 18 studies with 25 total comparisons from qualified studies that found a small positive tDCS effect on alcohol craving and bilateral tDCS significantly reducing alcohol craving9. Mostafavi, et al .2020 in a systemic review and meta-analysis study analyzed a total of 34 studies of which there are 11 tDCS trials found that there is no evidence of the positive effect of TMS/tDCS on various dimensions of Alcohol dependence. In our study, we are using 2 different areas i.e. DLPFC & Mid-cerebellum^10^.So the results of previous studies using prefrontal tDCS for ADS are ambiguous. Cerebellar tDCS had been tried in food cravings with inconclusive results. To our knowledge, this is the first study in ADS using Prefronto-cerebellar tDCS. We are using HD-tDCS because it provides a more focalized stimulation. We are adding the cerebellum as a novel site to reduce craving & relapse in ADS patients. By stimulating the cerebellum, we expect to improve the executive function & response inhibition which are important underlying mechanisms for continuing substance use. As discussed earlier treating relapse in ADS is a real challenge. This study will strengthen our understanding of the functionality & connectivity of the cerebellum. Neuromodulation in psychiatry is a growing science. Our study may be replicated in translational psychiatry & further enhance our understanding of tDCS & its application.

## CONCLUSION

This study will help to understand the effect of Prefronto-cerebellar tDCS in reducing relapse and craving in patients with ADS. It will enhance our current understanding of Cerebellar tDCS in addiction, especially in Alcohol Dependence.

## Data Availability

No datasets were generated or analysed during the current study. All relevant data from this study will be made available upon study completion

## CONFLICT OF INTEREST

None

## FUNDING

We got the Indian Council of Medical Research (ICMR) award for financial support for the PG-Thesis. [(No.3/2/June-2021/PG-Thesis-HRD(34)]

## Notes

### Competing Interest Statement

The authors have declared no competing interest.

### Clinical Trial

Clinical Trial Register India (CTRI) Registration No: CTRI/2021/09/036989. Reference no: REF/2021/05/043836

### Author Declarations

At the convened meeting of IEC-AIIMS Raipur held on 29.05.2021, the IEC voted to approve the above referenced protocol. letter by IEC Raipur

